# Global, Regional and National Incidence and Case-fatality rates of Novel Coronavirus (COVID-19) across 154 countries and territories: A systematic assessment of cases reported from January to March 16, 2020

**DOI:** 10.1101/2020.03.26.20044743

**Authors:** Akshaya Srikanth Bhagavathula, Jamal Rahmani, Wafa Ali Aldhaleei, Pavan Kumar, Alessandro Rovetta

## Abstract

**Background:** The 2019 novel coronavirus disease (COVID-19) outbreak turned into a pandemic, with hundreds of thousands of cases reported globally. The number of cases dramatically increased beginning in early March 2020.

**Aim:** We assessed the cumulative change in the incidence and case-fatality rates of COVID-19 at the global, regional, and national levels from January to March 16, 2020, in 154 affected countries and territories globally.

**Methods:** We collected data of COVID-19 cases using the GitHub repository, which provided real-time surveillance information developed by the Center for Systems Science and Engineering (CSSE), Johns Hopkins University (USA). Information such as confirmed COVID-19 cases, deaths, and recoveries reported across all affected countries was collected from January 22 to March 16, 2020. We estimated the change in the incidence rate, case-fatality rate, and recovery rate from January 22 to February 29 and from March 1 to March 16, 2020.

**Results:** From January 22 to March 16, 2020, globally, the number of incident COVID-19 cases increased by 276.2%, and Europe recorded 65,281 new cases from March 1 to 16, 2020. Overall, the case-fatality rate was 3.92%, with a high COVID-19 fatality rate in Italy (7.7%), Iran (5.7%), China (4.2%) and the United Kingdom (3.6%). The estimated percentage change in COVID-19 cases from March 1 to 16, 2020, was highest in Belgium (105.8/100,000 population), followed by Qatar (439/100,000 population) and Portugal (331/100,000 population). The overall recovery rate of COVID-19 was 43%; China (35.5%) had the highest recovery rate, while the United States of America recorded a recovery rate of 0.3%.

**Conclusion:** Overall, all the COVID-19-affected countries showed an upward trend in incidence, with little change in the incidence rate of -0.20% from January to Mid-March. The case-fatality rate was found to be 3.92%, and the recovery rate was observed to be less than half (43%) among COVID-19 patients. Italy, Iran, and Spain had the largest numbers of new cases of COVID-19 from March 1 to 16, 2020.

## Introduction

In early January, a cluster of pneumonia cases of unknown etiology from Wuhan, Hubei Province in China, was identified as novel coronavirus, which is now referred to as severe acute respiratory syndrome coronavirus 2 (SARS-CoV-2) and the illness as coronavirus disease 2019 (COVID-19) [1]. In January, COVID-19 spread throughout Wuhan with several cases, and other countries started to report their first cases of the disease. The outbreak of COVID-19 continues to spread around the world and has become a significant public health problem [2]. Subsequently, several hundreds of thousands of cases were reported around the world, and on March 11, 2020, the World Health Organization declared COVID-19 a pandemic [3].

As of March 31, 2020, over 800,000 cases of COVID-19 have been reported across 203 countries and territories, and more than 38,000 deaths had been reported [4]. Currently, the dynamics of COVID-19 are placing tremendous strain on countries, citizens, resources, and economies. In the absence of an antiviral treatment or vaccine against SARS-CoV-2, preventive measures such as hand hygiene practices, social distancing measures, movement restrictions, self-quarantines, closure of institutions, and other interventions have been intensified [5]. As these interventions are gradually implemented, early epidemiological estimates of COVID-19 distribution, cumulative changes in the incidence, and case-fatality rate are crucial for effective interventions. Therefore, we assessed the cumulative change in the incidence and case-fatality rates of COVID-19 at global, regional, and national levels from January 22 – March 16, 2020, across 154 affected countries and territories globally.

## Methods

In our analysis, we used the GitHub data repository, a publicly available dataset of the newly confirmed COVID-19 cases updated daily [6]. First, we extracted the data of confirmed cases, death, and recovery reported from January 22 to March 16, 2020. Second, the collected information was filtered and categorized into regions. These data were used to assess the trends in COVID-19 cases, case-fatality rates, and recovery rates at the global, regional, and national levels. Thus, we calculated the relative change in the incident cases and deaths reported in each country and territory and truncated percentage changes in the new cases and the case-fatality rate during the January 22 – March 16, 2020 period. We also reported the recovery rate of the known cases in each country.

### Statistical analysis

Frequencies and percentages are used to summarize the information. The case-fatality rate was defined as (number of deaths reported/total number of cases) × 100. The estimated percentage change was calculated using the formula: ((y2-y1)/y1)*100, where y1= initial value and y2= end value. All statistical analyses were conducted using STATA 16 data analysis and statistical software (StataCorp, LLC, TX, USA).

## Results

As of March 16, 2020, the global incidence of COVID-19 increased to 181,546 cases across 154 countries and territories, with a case-fatality rate of 3.92% and a recovery rate of 43%. The distribution of COVID-19 at the regional and national levels is presented in Table 1. For European and North American countries, the number of new cases showed an upward trend, and Europe reported a higher case-fatality rate (4.2%) than Asia (3.8%). By the middle of March, China (n=81033), Italy (n=27980), Iran (n=14991), Spain (n=9942), South Korea (n=8236) and Germany (n=7272) reported the highest numbers of COVID-19 cases. However, of these, Italy (7.7%), Iran (5.7%), and China (4.2%) reported a higher mortality rate than the global average (3.9%).

**Table 1:** Global distribution of COVID-19 cases, mortality, and recovery from until March 16, 2020.

| Characteristics | End of February, 2020 |  |  | Mid-March, 2020 |  |  | Case-fatality rate (%) | Cure rate (%) |
| --- | --- | --- | --- | --- | --- | --- | --- | --- |
|  | *Cases* | *Died* | *Recovered* | *Cases* | *Died* | *Recovered* |  |  |
| **Global** | 86012 | 2941 | 39782 | 181,546 | 7126 | 78088 | 3.92 | 43 |
| **Region** |  |  |  |  |  |  |  |  |
| Asia | 83715 | 2903 | 39659 | 108376 | 4207 | 74190 | 3.88 | 68.4 |
| Africa | 3 | 0 | 1 | 411 | 8 | 42 | 1.94 | 10.2 |
| Europe | 1467 | 31 | 88 | 65748 | 2803 | 3474 | 4.2 | 5.2 |
| Australia | 25 | 0 | 11 | 377 | 3 | 23 | 0.8 | 6.1 |
| North America | 92 | 1 | 13 | 5277 | 93 | 32 | 1.76 | 0.6 |
| South America | 2 | 0 | 0 | 650 | 5 | 2 | 0.76 |  |
| Oceanic | 1 | 0 | 0 | 11 | 0 | 0 | - |  |
| Cruise ship | 705 | 6 | 10 | 696 | 7 | 325 | 1 | 47 |
| **Asia** |  |  |  |  |  |  |  |  |
| China | 79356 | 2837 | 39320 | 81033 | 3430 | 28804 | 4.23 | 35.5 |
| Thailand | 42 | 43 | 123 | 147 | 1 | 12 | 0.68 | 8.1 |
| Japan | 241 | 16 | 27 | 825 | 27 | 113 | 3.27 | 13.7 |
| Singapore | 102 | 5 | 32 | 243 | 0 | 37 | - | 15.2 |
| Korea, South | 3150 | 0 | 18 | 8236 | 75 | 1110 | 0.91 | 13.4 |
| Taiwan* | 39 | 0 | 0 | 67 | 1 | 11 | 1.49 | 16.4 |
| Malaysia | 25 | 0 | 1 | 566 | 0 | 24 | - | 4.2 |
| United Arab Emirates | 21 | 0 | 72 | 98 | 0 | 18 | - | 18.4 |
| Vietnam | 16 | 0 | 0 | 61 | 0 | - | - | - |
| Cambodia | 1 | 0 | 28 | 7 | 0 | - | - | - |
| India | 3 | 1 | 1 | 119 | 2 | 10 | 1.68 | 8.4 |
| Nepal | 1 | 0 | 0 | 1 | 0 | - | - | - |
| Philippines | 3 | 0 | 0 | 142 | 12 | 1 | 8.45 | 0.7 |
| Sri Lanka | 1 | 0 | 0 | 28 | 0 | - | - | - |
| Afghanistan | 1 | 0 | 0 | 21 | 0 | 1 | - | 4.8 |
| Bahrain | 41 | 0 | 3 | 214 | 1 | 77 | 0.47 | 36.0 |
| Bangladesh | 0 | 0 | 0 | 8 | 0 | 2 | - | 25.0 |
| Brunei | 0 | 0 | 0 | 54 | 0 | - | - | - |
| Indonesia | 0 | 0 | 5 | 134 | 5 | 8 | 3.73 | 6.0 |
| Iran | 593 | 1 | 9 | 14991 | 853 | 4467 | 5.69 | 29.8 |
| Iraq | 13 | 0 | - | 124 | 10 | 26 | 8.06 | 21.0 |
| Israel | 7 | 0 | - | 255 | 0 | 3 | - | 1.2 |
| Jordan | 0 | 0 | - | 17 | 0 | 1 | - | 5.8 |
| Kazakhstan | 0 | 0 | 1 | 10 | 0 | - | - | - |
| Kenya | 0 | 0 | 0 | 3 | 0 | - | - | - |
| Kuwait | 45 | 0 | - | 123 | 0 | 9 | - | 7.3 |
| Lebanon | 4 | 0 | 0 | 99 | 3 | 1 | 3.03 | 1 |
| Maldives | 0 | 0 | - | 13 | 0 | - |  | - |
| Mongolia | 0 | 0 | - | 1 | 0 | - |  | - |
| Palestinian | 0 | 0 | 0 | 1 | 0 | - |  | - |
| Oman | 6 | 0 | - | 22 | 0 | 8 |  | 36.3 |
| Pakistan | 4 | 0 | - | 136 | 0 | 2 |  | 1.4 |
| Qatar | 1 | 0 | - | 439 | 0 | 4 |  | 0.9 |
| Saudi Arabia | 0 | 0 | - | 118 | 0 | 2 |  | 1.7 |
| Turkey | 0 | 0 | - | 18 | 0 | - |  | - |
| Uzbekistan | 0 | 0 | - | 6 | 0 | - |  | - |
| **Europe** |  |  |  |  |  |  |  |  |
| Italy | 1128 | 29 | 46 | 27980 | 2158 | 2703 | 7.71 | 9.6 |
| Spain | 45 | 0 | 2 | 9942 | 342 | 528 | 3.44 | 5.3 |
| Germany | 79 | 0 | 0 | 7272 | 17 | - | 0.23 | - |
| France | 100 | 2 | 0 | 6650 | 148 | - | 2.23 | - |
| Switzerland | 18 | 0 | 0 | 2200 | 14 | 4 | 0.64 | 0.2 |
| United Kingdom | 23 | 0 | 8 | 1551 | 56 | 13 | 3.61 | 0.8 |
| Netherlands | 6 | 0 | 0 | 1414 | 24 | 2 | 1.70 | 0.1 |
| Norway | 15 | 0 | 0 | 1333 | 3 | 1 | 0.22 | 0.07 |
| Sweden | 12 | 0 | 0 | 1103 | 6 | 1 | 0.54 | 0.09 |
| Belgium | 1 | 0 | - | 1058 | 5 | - | 0.47 | - |
| Austria | 9 | 0 | 0 | 1018 | 3 | 6 | 0.29 | 0.6 |
| Denmark | 3 | 0 | 0 | 932 | 3 | 1 | 0.32 | 0.1 |
| Greece | 4 | 0 | 0 | 331 | 4 | 8 | 1.21 | 2.4 |
| Portugal | 0 | 0 | 0 | 331 | 0 | 3 | - | 0.9 |
| Czechia | 0 | 0 | 0 | 298 | 0 | 3 | - | 1 |
| Finland | 3 | 0 | 1 | 277 | 0 | 9 | - | 3.2 |
| Slovenia | 0 | 0 | - | 253 | 1 | - | 0.40 | - |
| Estonia | 1 | 0 | 0 | 205 | 0 | 1 | - | 0.5 |
| Iceland | 1 | 0 | - | 180 | 0 | - | - | - |
| Poland | 0 | 0 | 0 | 177 | 4 | 13 | 2.26 | 7.3 |
| Ireland | 1 | 0 | - | 169 | 2 | - | 1.18 | - |
| Romania | 3 | 0 | 0 | 158 | 0 | 9 | - | 5.7 |
| San Marino | 1 | 0 | 0 | 109 | 7 | 4 | 6.42 | 3.7 |
| Russia | 2 | 0 | 2 | 90 | 0 | 6 | - | 6.6 |
| Luxembourg | 1 | 0 | - | 77 | 1 | - | 1.30 | - |
| Slovakia | 0 | 0 | - | 63 | 0 | - | - | - |
| Croatia | 6 | 0 | 0 | 57 | 0 | 2 | - | 3.5 |
| Serbia | 0 | 0 | 0 | 55 | 0 | 1 | - | 1.8 |
| Armenia | 0 | 0 | - | 52 | 0 | - | - | - |
| Bulgaria | 0 | 0 | - | 52 | 2 | - | 3.85 | - |
| Albania | 0 | 0 | - | 51 | 1 | - | 1.96 | - |
| Hungary | 0 | 0 | 0 | 39 | 1 | 1 | 2.56 | 2.5 |
| Belarus | 1 | 0 | 0 | 36 | 0 | 3 | - | 8.3 |
| Latvia | 0 | 0 | 0 | 34 | 0 | 1 | - | 2.9 |
| Cyprus | 0 | 0 | - | 33 | 0 | - | - | - |
| Georgia | 1 | 0 | 16 | 33 | 0 | 1 | - | 3 |
| Malta | 0 | 0 | 0 | 30 | 0 | 2 | - | 6.6 |
| Bosnia and Herzegovina | 0 | 0 | - | 25 | 0 | - | - | - |
| Moldova | 0 | 0 | - | 23 | 0 | - | - | - |
| North Macedonia | 1 | 0 | 0 | 18 | 0 | 1 | - | 5.5 |
| Lithuania | 1 | 0 | 0 | 17 | 0 | 1 | - | 5.8 |
| Monaco | 1 | 0 | - | 7 | 0 | - | - | - |
| Ukraine | 0 | 0 | - | 7 | 1 | - | 14.2 | - |
| Liechtenstein | 0 | 0 | - | 4 | 0 | - | - | - |
| Andorra | 0 | 0 | 0 | 2 | 0 | 1 | - | 50 |
| Guernsey | 0 | 0 | - | 1 | 0 | - | - | - |
| Holy See | 0 | 0 | - | 1 | 0 | - | - | - |
| **North America** |  |  |  |  |  |  |  |  |
| United States of America | 68 | 1 | 7 | 4632 | 85 | 17 | 1.83 | 0.3 |
| Canada | 20 | 0 | 6 | 415 | 4 | 9 | 0.96 | 2.1 |
| Panama | 0 | 0 | - | 55 | 1 | - | 1.81 | - |
| Mexico | 4 | 0 | - | 53 | 0 | 4 | 0 | 7.5 |
| Costa Rica | 0 | 0 | - | 35 | 0 | - | 0 | - |
| Martinique | 0 | 0 | - | 15 | 1 | - | 6.66 | - |
| Dominican Republic | 0 | 0 | - | 11 | 0 | - | 0 | - |
| Jamaica | 0 | 0 | - | 10 | 0 | 2 | 0 | 20 |
| Guadeloupe | 0 | 0 | - | 6 | 0 | - | 0 | - |
| Honduras | 0 | 0 | - | 6 | 0 | - | 0 | - |
| Cuba | 0 | 0 | - | 4 | 0 | - | 0 | - |
| Trinidad and Tobago | 0 | 0 | - | 4 | 0 | - | 0 | - |
| Puerto Rico | 0 | 0 | - | 3 | 0 | - | 0 | - |
| Guatemala | 0 | 0 | - | 2 | 1 | - | 50 | - |
| Jersey | 0 | 0 | - | 2 | 0 | - | 0 | - |
| Kosovo | 0 | 0 | - | 2 | 0 | - | 0 | - |
| Saint Lucia | 0 | 0 | - | 2 | 0 | - | 0 | - |
| Antigua and Barbuda | 0 | 0 | - | 1 | 0 | - | 0 | - |
| Greenland | 0 | 0 | - | 1 | 0 | - | 0 | - |
| Liberia | 0 | 0 | - | 1 | 0 | - | 0 | - |
| Saint Vincent | 0 | 0 | - | 1 | 0 | - | 0 | - |
| and the<br>Grenadines |  |  |  |  |  |  |  |  |
| The Bahamas | 0 | 0 | - | 1 | 0 | - | 0 | - |
| **South America** |  |  |  |  |  |  |  |  |
| Brazil | 2 | - | - | 200 | 0 | 1 | - | 0.5 |
| Argentina | 0 | - | - | 56 | 2 | 1 | 3.57 | 1.7 |
| Aruba | 0 | - | - | 2 | 0 | - | - | - |
| Bolivia | 0 | - | - | 11 | 0 | - | - | - |
| Chile | 0 | - | - | 155 | 0 | - | - | - |
| Colombia | 0 | - | - | 54 | 0 | - | - | - |
| Ecuador | 0 | - | - | 37 | 2 | - | 5.40 | - |
| French Guiana | 0 | - | - | 11 | 0 | - | - | - |
| Guyana | 0 | - | - | 4 | 1 | - | 25 | - |
| Paraguay | 0 | - | - | 8 | 0 | - | - | - |
| Peru | 0 | - | - | 86 | 0 | - | - | - |
| Suriname | 0 | - | - | 1 | 0 | - | - | - |
| Uruguay | 0 | - | - | 8 | 0 | - | - | - |
| Venezuela | 0 | - | - | 17 | 0 | - | - | - |
| **Africa** |  |  |  |  |  |  |  |  |
| Algeria | 1 | - | 0 | 54 | 4 | 12 | 7.4 | 22.2 |
| Egypt | 1 | - | 1 | 150 | 2 | 27 | 1.3 | 18 |
| Nigeria | 1 | - | - | 2 | 0 | - | - | - |
| Benin | 0 | - | - | 1 | 0 | - | - | - |
| Burkina Faso | 0 | - | - | 15 | 0 | - | - | - |
| Cameroon | 0 | - | - | 4 | 0 | - | - | - |
| Central African<br>Republic | 0 | - | - | 1 | 0 | - | - | - |
| Congo | 0 | - | - | 3 | 0 | - | - | - |
| Cote d'Ivoire | 0 | - | - | 1 | 0 | - | - | - |
| Equatorial<br>Guinea | 0 | - | - | 1 | 0 | - | - | - |
| Eswatini | 0 | - | - | 1 | 0 | - | - | - |
| Ethiopia | 0 | - | - | 5 | 0 | - | - | - |
| Gabon | 0 | - | - | 1 | 0 | 1 | - | 100 |
| Ghana | 0 | - | - | 6 | 0 | - | - | - |
| Guinea | 0 | - | - | 1 | 0 | - | - | - |
| Mauritania | 0 | - | - | 1 | 0 | - | - | - |
| Mayotte | 0 | - | - | 1 | 0 | - | - | - |
| Morocco | 0 | - | - | 29 | 1 | 1 | 3.4 | 3.4 |
| Namibia | 0 | - | - | 2 | 0 | - | - | - |
| Republic of the<br>Congo | 0 | - | - | 1 | 0 | - | - | - |
| Reunion | 0 | - | - | 9 | 0 | - | - | - |
| Rwanda | 0 | - | - | 5 | 0 | - | - | - |
| Senegal | 0 | - | - | 24 | 0 | 2 | - | 8.3 |
| Seychelles | 0 | - | - | 3 | 0 | - | - | - |
| Somalia | 0 | - | - | 1 | 0 | - | - | - |
| South Africa | 0 | - | - | 62 | 0 | - | - | - |
| Sudan | 0 | - | - | 1 | 1 | - | 100 | - |
| Tanzania | 0 | - | - | 1 | 0 | - | - | - |
| Togo | 0 | - | - | 1 | 0 | - | - | - |
| Tunisia | 0 | - | - | 20 | 0 | - | - | - |
| **Australia** | 16 | - | 9 | 377 | 3 | 14 | 0.8 | 3.7 |
| New Zealand | 1 | - | - | 8 | - | - | - | - |
| Guam | 0 | - | - | 3 | - | - | - | - |
| **Cruise ship** | 705 | 6 | 10 | 696 | 7 | 325 | 1 | 46.7 |

From January to March 16, 2020, the COVID-19 incidence increased in a total of 154 countries (Figure 1A). Within a short period, the COVID-19 cases dramatically increased all over the world, except for in some sub-Saharan African countries. From March 1 to 16, 2020, the number of incident COVID-19 cases increased in a total of 152 countries (Figure 1B). The percent change in the incidence cases was less than 1000% in 70 countries, including China (102%), while the greatest increase in percentage change was observed in Belgium (1058/100,000 population), Qatar (439/100,000 population), Portugal (33,100%), and Denmark (310.6/100,000 population).

**Figure 1:**
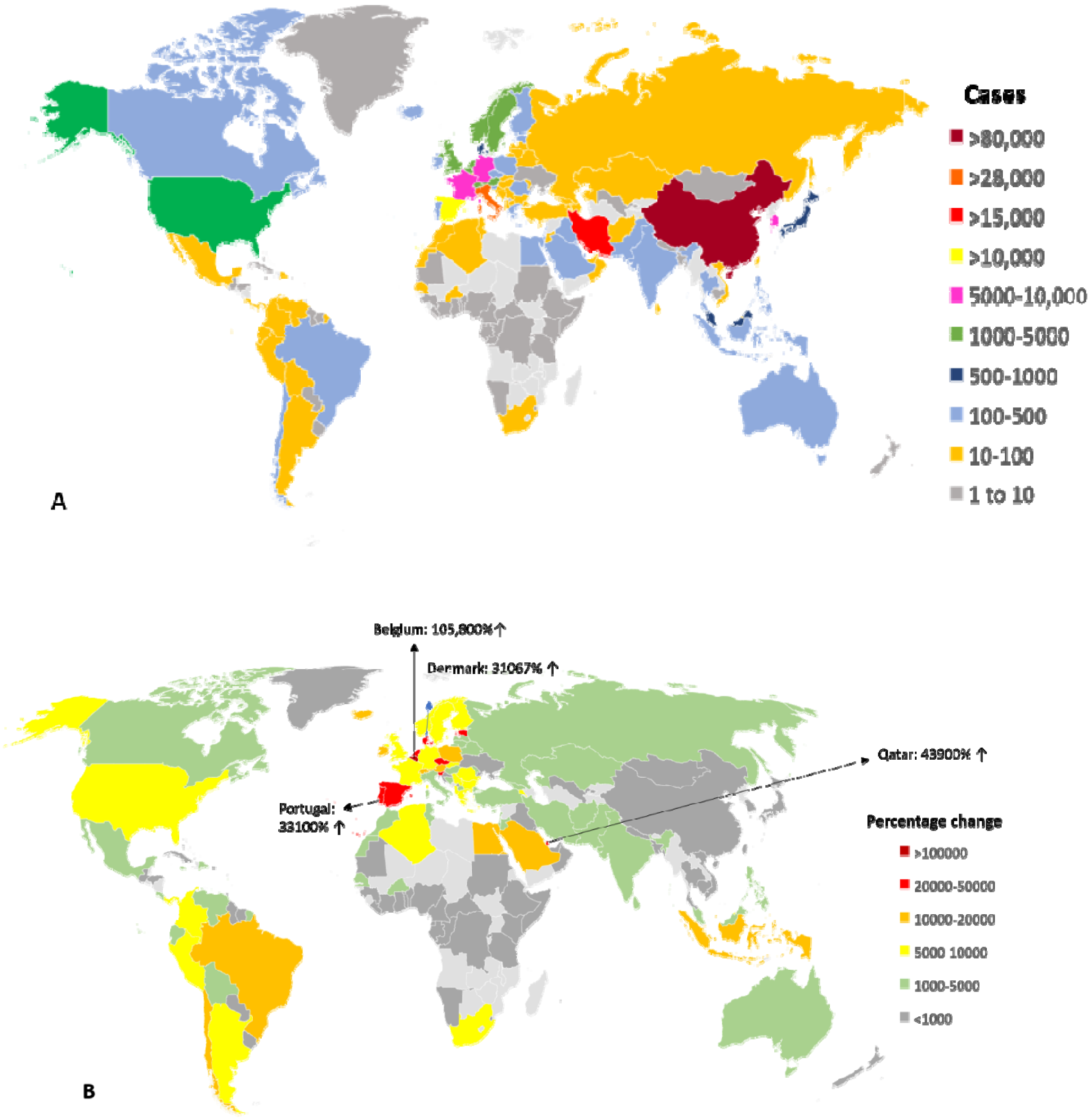
The global burden of COVID-19 across 154 countries January to March 16, 2020. (A) Total number of cases reported; (B) The relative change in incident cases from March 1 to 16, 2020.

Globally, from January to March 16, 2020, the number of new COVID-19 cases worldwide increased by 276.2% from 555 cases to 153,312 [Table 2]. With a decline in the number of new cases in China, a slight change in the incidence of -0.20% was noticed during the first half of March 2020. However, the most pronounced increase in new COVID-19 cases was observed in Europe by 66,749%, from “0” cases in January to 1467 cases in February to 65,281 cases in mid-

**Table 2:** Temporal trends in the COVID-19 pandemic from January-March 16, 2020.

| Characteristics | Through January 26, 2020 | New cases identified through February 29, 2020 | Change in incidence (%) | New cases identified from March 1 to 16, 2020 | Change in incidence | Overall incidence change from January to Mid-March (%) |
| --- | --- | --- | --- | --- | --- | --- |
| Global | 555 | 85457 | 152.9 | 67855 | -0.20 | 276.2 |
| **Region** |  |  |  |  |  |  |
| Asia | 554 | 83161 | 150.1 | 24,664 | -0.70 | 193.6 |
| Africa | 0 | 3 | 3 | 404 | 133.6 | 407 |
| Europe | 0 | 1467 | 1467 | 65,281 | 42.8 | 66,749 |
| Australia | 0 | 25 | 25 | 352 | 13 | 378 |
| North America | 1 | 92 | 91 | 5187 | 56 | 5278 |
| South America | 0 | 2 | 2 | 648 | 323 | 650 |
| Oceanic | 0 | 1 | 1 | 10 | 9 | 10 |
| Cruise ship | 0 | 705 | 705 | -9 | -1.01 | 696 |
| **Asia** |  |  |  |  |  |  |
| China | 548 | 78808 | 142.8 | 2225 | -0.97 | 3.1 |
| Iran | 0 | 593 | 593 | 14398 | 23.28 | 593 |
| South Korea | 1 | 3149 | 3148 | 5087 | 0.62 | 5086 |
| Japan | 2 | 239 | 118.5 | 586 | 1.45 | 292 |
| Malaysia | 0 | 25 | 25 | 541 | 20.64 | 25 |
| Qatar | 0 | 1 | 1 | 438 | 437 | 1 |
| Israel | 0 | 7 | 7 | 248 | 34.43 | 7 |
| Bahrain | 0 | 41 | 41 | 173 | 3.22 | 41 |
| Singapore | 0 | 102 | 102 | 141 | 0.38 | 102 |
| Philippines | 0 | 3 | 3 | 139 | 45.33 | 3 |
| Indonesia | 0 | 0 | 0 | 134 | 134 | 0 |
| Pakistan | 0 | 4 | 4 | 132 | 32 | 4 |
| Saudi Arabia | 0 | 0 | 0 | 118 | 118 | 0 |
| India | 0 | 3 | 3 | 116 | 37.67 | 3 |
| Iraq | 0 | 13 | 13 | 111 | 7.54 | 13 |
| Thailand | 2 | 40 | 19 | 107 | 1.68 | 52.5 |
| **Europe** |  |  |  |  |  |  |
| Italy | 0 | 1128 | 1128 | 27980 | 22.80 | 27980 |
| Spain | 0 | 45 | 45 | 9942 | 218.93 | 9942 |
| Germany | 0 | 79 | 79 | 7272 | 90 | 7272 |
| France | 0 | 100 | 100 | 6650 | 64.50 | 6650 |
| Switzerland | 0 | 18 | 18 | 2200 | 120.22 | 2200 |
| United Kingdom | 0 | 23 | 23 | 1551 | 65.43 | 1551 |
| Netherlands | 0 | 6 | 6 | 1414 | 233.67 | 1414 |
| Norway | 0 | 15 | 15 | 1333 | 86.87 | 1333 |
| Sweden | 0 | 12 | 12 | 1103 | 89.92 | 1103 |
| Belgium | 0 | 1 | 1 | 1058 | 1056.00 | 1058 |
| Austria | 0 | 9 | 9 | 1018 | 111.11 | 1018 |
| Denmark | 0 | 3 | 3 | 932 | 308.67 | 932 |
| Greece | 0 | 4 | 4 | 331 | 80.75 | 331 |
| Portugal | 0 | 0 | 0 | 331 | 33 | 331 |
| Czechia | 0 | 0 | 0 | 298 | 298 | 298 |
| Finland | 0 | 3 | 3 | 277 | 90.33 | 277 |
| Slovenia | 0 | 0 | 0 | 253 | 253 | 253 |
| Estonia | 0 | 1 | 1 | 205 | 203 | 205 |
| Iceland | 0 | 1 | 1 | 180 | 178 | 180 |
| Poland | 0 | 0 | 0 | 177 | 177 | 177 |
| Ireland | 0 | 1 | 1 | 169 | 167 | 169 |
| Romania | 0 | 3 | 3 | 158 | 50.67 | 158 |
| San Marino | 0 | 1 | 1 | 109 | 107 | 109 |
| **North America** |  |  |  |  |  |  |
| United States of America | 1 | 67 | 66 | 4612 | 67.83 | 4679 |
| Canada | 0 | 20 | 20 | 395 | 18.7 | 415 |
| **South America** |  |  |  |  |  |  |
| Brazil | 0 | 2 | 2 | 148 | 73 | 151 |
| Chile | 0 | 0 | 0 | 155 | 155 | 155 |
| **Africa** |  |  |  |  |  |  |
| Egypt | 0 | 1 | 1 | 149 | 148 | 149 |

March 2020. The affected countries and territories experienced an increase in incidence, except the cases in China, which had an incidence of -0.97%, and the cases reported from a cruise ship (-1.01%). In March, South Korea (0.62%), Singapore (0.38%), and Thailand (1.68%) showed a decrease in COVID-19 incidence rates compared to the previous period.

A total of 7126 COVID-19 deaths occurred globally as of March 16, 2020, with a relative change of 242.2% compared to the deaths that occurred through the end of February 2020 (Figure 2A). From March 1 to 16, a relative increase in the death rate was observed in Iran (8.53%) and Spain (3.42%) (Figure 2B).

**Figure 2:**
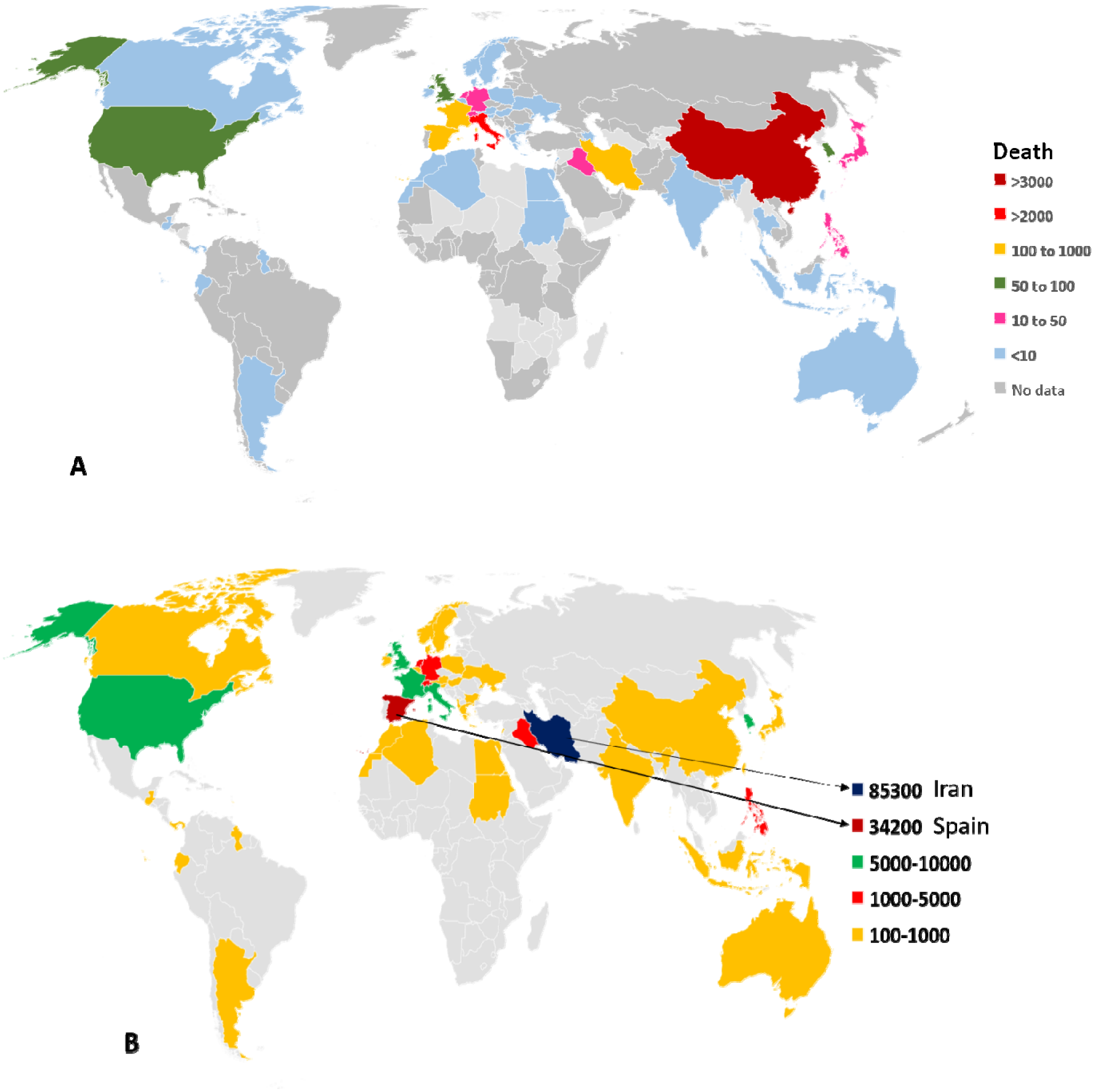
Global mortality associated with COVID-19 pandemic from between January to March 16, 2020. (A) Total number of deaths reported and (B) The relative change in incident deaths (EPC: estimated percentage change) March 1 to 16, 2020.

## Discussion

Between January 22 and March 16, 2020, the number of COVID-19 cases increased rapidly worldwide, and thousands of deaths have been reported in several countries. Our results indicate that the global trends in the ongoing COVID-19 pandemic rapidly emerged in all countries in early March 2020, and the observed incidence increased by 276.2%. In particular, an increase in incidence and mortality rates were prominent in most regions worldwide (albeit the majority of cases still occurred in China and Italy). Nevertheless, the incidence of COVID-19 cases varies significantly between countries and regions. The difference between the highest and the lowest incidence was 1088-fold (Belgium: 1056 versus China: 0.97), and the difference in the mortality rate was 26.6-fold (Italy: 7.71 versus Austria: 0.29). It is worth noting that countries such as South Korea, Singapore, and Thailand showed a decreasing trend in the COVID-19 incidence rates in early March 2020.

Our estimates of the case-fatality rate of COVID-19 in Italy and Iran are nearly double that of the global case-fatality rate (3.9%). These findings reflect the situation in these countries. This suggests that the situation of COVID-19 in these countries is particularly dire and requires intensive support to reduce fatalities. Moreover, insufficient healthcare resources in many countries, inadequate screening among travelers, lack of preparedness of healthcare workers, and misconceptions among the general public can lead to substantial diagnosis gaps, which may have caused the disproportionate distribution in incidence and mortality. Our data are in agreement with the estimates of the worldwide incidence and mortality from the World health organization (WHO), the United States Centers for Disease Control and Prevention (CDC), and the European Center for Disease Prevention and Control (ECDC) [7-9]. Regarding the regional variations in the incidence of COVID-19 from March 1 to 16, the highest values were found in South America and Africa and the lowest were found in Australia and parts of Asia.

Improvement in early screening, diagnosis, and strict adherence to preventive measures may play a key role in halting the global epidemiological transition, including the numbers of incident cases and deaths. These factors might have been responsible for the sharp changes observed in China, Singapore, Hong Kong, Taiwan, and South Korea [10-12]. Indeed, mainland in China and neighboring countries experienced a sharp increase in incidence, as well as in death, in February 2020. The following factors may have contributed to the gradual leveling off in incidence: a) responding aggressively by implementing travel restrictions on passengers to prevent transmission [13], b) early recognition and epidemic preparedness with 124 “action items,” including border control, school closures, and work policies [14] and (c) rigorous detection and strict quarantine measures to break the long chain of COVID-19 outbreaks [15]. However, further epidemiological analysis addressing the current situation, knowledge gaps in its etiology, and comprehensive management of COVID-19 at national and regional levels are warranted.

The study has some unavoidable limitations. Our analysis relies on epidemiologic data reported through March 16, 2020. Therefore, the accuracy of the results depends on the quality of the data reported. In terms of quality, the Center for Systems Science and Engineering (CSSE) at John Hopkins University (USA) obtained confirmed information from respective centers prior to the collection of information and is coordinated by a team of experts from John Hopkins University [16]. Most of the information presented in the GitHub repository by the CSSE is consistent with the data reported by the WHO [16]. There may be missing information due to limited reporting resources, delays in investigations, and reluctance to update the information in some countries. Thus, information bias is inevitable. Due to the limitations of the data, we cannot perform further investigation in regard to the clinical, etiological, treatment and risk stratification of the COVID-19 cases.

### Conclusion

Overall, all the COVID-19-affected countries showed an upward trend in incidence globally, with little change in the incidence rate of -0.20% from January to Mid-March. The case-fatality rate was found to be 3.92%, and the recovery rate was observed to be less than half (43%) among COVID-19 patients. Italy, Iran, and Spain had most new cases of COVID-19 from March 1 to 16, 2020.

## Data Availability

Data is readily available upon contacting the corresponding author.

## Acknowledgment

We thank all the John Hopkins institute, USA for providing the data study participants for their voluntary participation and for providing essential information.

## Authors’ contributions

ASB, JR designed the study, developed the study, collected the data, analyzed the data, and prepared the manuscript. WAA, PK and AR designed the study tools, conducted the analysis, and conducted the literature review. ASB, WAA, PK and JR filtered and analyzed the data. All authors read and approved the final manuscript.

## Available data and materials

All data obtained through data respiratory are readily available and can be contacted to corresponding author.

## Consent for publication

Not applicable.

